# Lack of validated blood pressure devices for use in pregnancy available from Australian pharmacies

**DOI:** 10.1101/2025.03.25.25324644

**Authors:** Kaylee Slater, Hollie Speer, Niamh Chapman, Dean S. Picone

## Abstract

Blood pressure monitoring is a critical aspect of prenatal care, as hypertension during pregnancy can lead to serious complications such as preeclampsia, eclampsia, and other hypertensive disorders. Automatic blood pressure devices are widely used for home monitoring due to their convenience and ease of use. Automatic blood pressure devices require additional accuracy validation before they are recommended for use in pregnancy. This study evaluated the availability of such devices from 18 Australian pharmacies. Only four devices (4/54, 7%) were validated for pregnancy and were more expensive than devices validated for the general population (14/54, 26%) and non-validated devices (40/54, 74%). Additionally, limited labelling and information was available to assist consumers to make informed purchasing decisions about home blood pressure devices for use in pregnancy. Increased availability, clear labelling and consumer education could help ensure use of appropriate blood pressure devices in pregnancy.

## Introduction

Blood pressure (BP) monitoring is a critical aspect of prenatal care, as hypertension during pregnancy can lead to serious complications such as preeclampsia, eclampsia, and other hypertensive disorders (1, 2). Automatic blood pressure devices are widely used for home monitoring due to their convenience and ease of use (3).

However, only 20% of home BP devices available on the market have been validated for use in general adult populations (4). To confirm accuracy among pregnant women, automated devices should undergo additional validation testing because of haemodynamic changes induced by pregnancy which may affect the accuracy of the algorithms used by automated oscillometric devices to measure BP (2, 5). Therefore, even fewer validated devices are available for use in this population (2). A systematic review of validation studies on office, ambulatory, and home BP measurement devices for pregnant women (6) found that out of 28 devices, 61% passed the validation criteria. However, 66% of these studies were conducted with protocol violations which raises questions about the reliability of the findings (6).

A much larger proportion of BP devices have been validated for general populations than for use during pregnancy. However, the real-world availability of home BP monitors that are validated for use during pregnancy is unknown. In Australia, most consumers purchase home BP devices from pharmacies (7), where the availability and validation status of such devices can vary significantly (4). Therefore, the aim of this study was to determine the availability of home BP devices validated for pregnancy from pharmacies, including device costs and claims regarding use in pregnancy.

## Methods

Pharmacies with physical and online stores in Australia selling upper-arm or wrist cuff BP devices were identified and searched by two independent reviewers (KS/HS) in May-June 2024. The search was conducted on Google Australia using the term “home blood pressure monitor/device”. Validation of devices specifically for use in pregnancy, and the general adult population was determined using the non-profit, hypertension expert-led STRIDE-BP database (8), which provides a rigorously peer-reviewed list of BP devices that have passed accuracy validation testing. Reviewers extracted the device cost and claims about use in pregnancy from instruction manuals and information on pharmacy webpages. Conflicts were adjudicated by other investigators (NC/DP).

## Results

Eighteen pharmacies and 54 unique home BP devices were identified. Four (7%) devices were validated for use in pregnancy and 14 (26%) were validated for use in the general population (Table 1). Sixteen out of 18 pharmacies stocked at least one device that was validated for use in pregnancy (Figure 1A).

**Table 1.** Home BP devices sold in 18 Australian pharmacies.

|  | **Number of**<br>**unique**<br>**devices found** | **Validated for use**<br>**in pregnancy, n**<br>**(%)** | **Validated for use**<br>**in general adult**<br>**population, n**<br>**(%)** | **No evidence of**<br>**validation in**<br>**general population**<br>**or pregnancy, n**<br>**(%)** |
| --- | --- | --- | --- | --- |
| **Upper**<br>**arm cuff** | 49 | 4 (8) | 13 (27) | 36 (74) |
| **Wrist**<br>**cuff** | 5 | 0 (0) | 1 (20) | 4 (80) |
| **Total** | 54 | 4 (7) | 14 (26) | 40 (74) |

BP: blood pressure. The percentages add to more than 100% because some devices validated in the general population were also validated for pregnancy.

**Figure 1.**
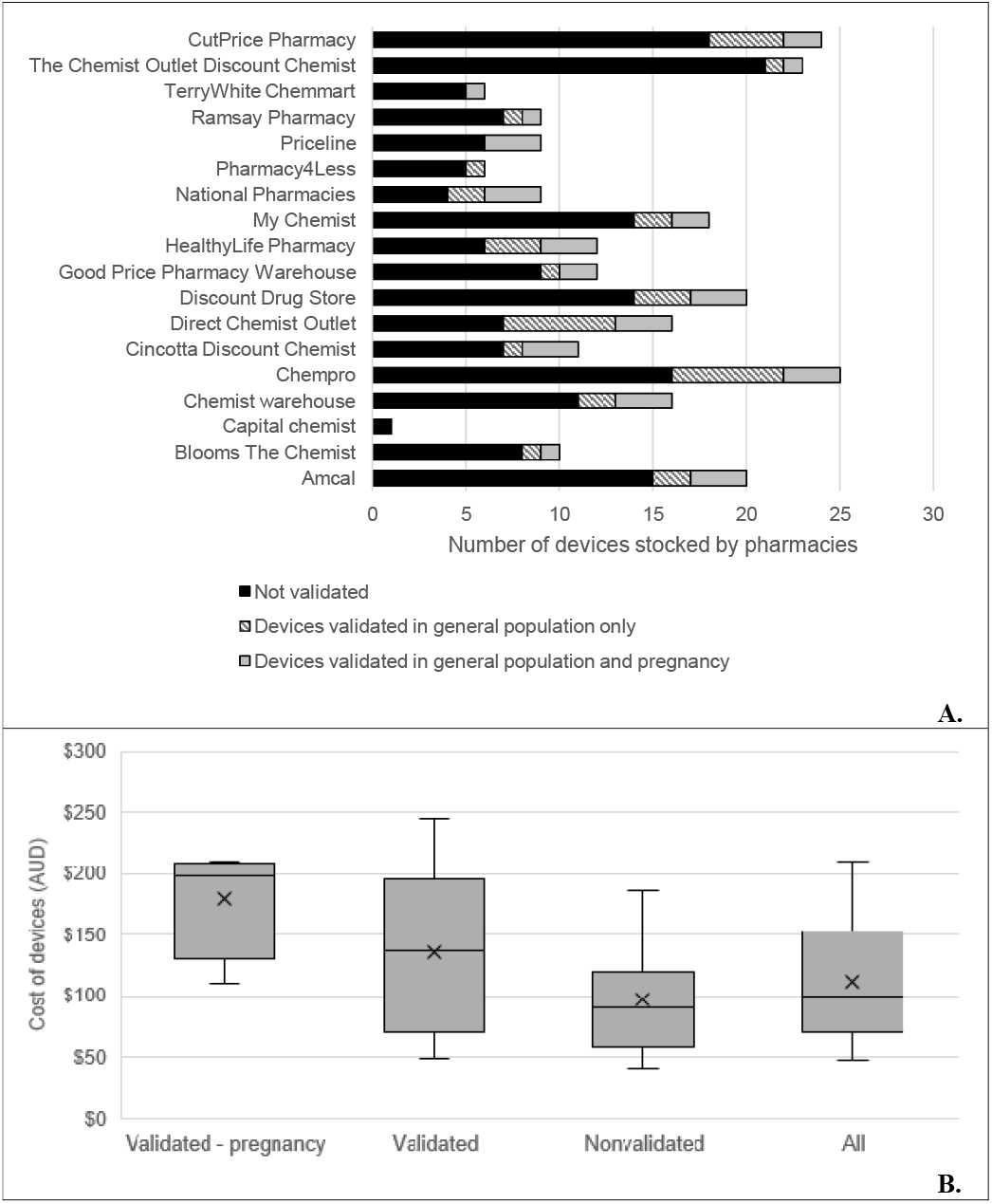
Availability of validated and non-validated devices from 18 Australian pharmacies (A, top) and cost of the home BP devices that are validated for use in pregnancy, validated in the general adult population, non validated and all combined (B, bottom). BP: blood pressure. B) Cost displayed in Australian dollars (12). Cost could not be determined for one device due to it only being available in-store. Devices labelled ‘validated’ also include those in the validated-pregnancy group. The ‘x’ on the plots represent the mean cost, middle line represents the median cost, the bottom edge of the box is Quartile 1 (value below which 25% of the data points fall) and the top edge is Quartile 3 (75%), the lowest point at the end of the lower whisker represents the lowest cost, and the upper edge of the upper whisker represents the highest cost.

Devices validated for use in pregnancy were more expensive than those validated in the general population (median AUD $198.99, interquartile range (IQR) 151.47-207.50 versus $137.47, IQR 69.99-192.99). Devices that had no evidence of validation were less expensive ($89.99, IQR 58.95-119.99) (Figure 1B).

Sixteen (30%) of the 54 devices listed a caution warning, suggesting users consult a healthcare provider before using the device in pregnancy. Of these devices, information about four stated they had not been tested among pregnant women and were not suitable for use.

This information was only available in device manuals and not at point-of-sale. Five devices stated they were recommended for use in pregnancy, although only one was validated. There was no information identified regarding use in pregnancy for 33 (61%) devices, although one of these devices was validated.

## Discussion

This study reveals a significant gap in the availability of validated BP devices for pregnant women in Australian pharmacies. Out of 54 home BP devices found in Australian pharmacies, only four were specifically validated for use in pregnancy and these devices were substantially more expensive than other devices. Although most pharmacies stocked at least one device that had been validated for use in pregnancy, there was limited information to assist consumers in making informed purchasing decisions at point of sale. Improved labelling and stakeholder education is needed to ensure the use of accuracy validated devices during pregnancy in Australia.

A recent systematic review and meta-analysis on the safety and efficacy of home BP monitoring during pregnancy found that home BP monitoring reduced hospital admissions by 70% and lowered the incidence of preeclampsia by 50%, compared to usual care (9). Nine studies were included in the meta-analysis, however, the authors noted concern about significant heterogeneity and quality of the evidence. Moreover, all the studies were conducted in Europe, United Kingdom and the United States of America, potentially limiting generalizability to other world regions and diverse populations. Indeed, the Society of Obstetric Medicine Australia and New Zealand guidelines (1) state that pregnant women with hypertension might benefit from home BP monitoring with a validated device, but that a current research priority for Australia is more locally relevant data on the utility and safety of home BP monitoring in pregnancy (1).

Despite the potential health benefits of home BP monitoring in pregnancy (10), there is minimal practical guidance for consumers to help them select a validated device, and for healthcare providers when advising patients (2, 4). A recent real-world study among 127 pregnant women from Australia (10) identified 40 different home BP devices being used by the participants, and although 20% were validated for use in the general population, none were specifically validated for use in pregnancy. This limited use of validated devices presents a challenge for pregnant women who need accurate home BP monitoring. In addition to practical guidance, the substantially higher cost of devices validated for use in pregnancy has the potential to influence consumer purchasing decisions. The lack of clear labelling and information on the validation status of these devices further complicates the purchasing process, which highlights the need for increased education and awareness among consumers and healthcare providers about the importance of using validated BP devices during pregnancy.

A limitation of this study is that data collection was restricted to Australian pharmacies with physical and online stores. Many online businesses stock home BP devices and previous work from Australia showed only 16% of upper-arm cuff devices from these sources were validated for use in the general population (4). However, a 2024 mixed-methods study found that most adults who measure their BP at home in Australia purchase their devices through pharmacies (7). Future research might explore global availability of validated home BP devices for pregnancy, including from e-commerce stores, such as Amazon. Moreover, the generalizability of the findings beyond Australia is unknown. However, previous work on the availability of validated BP devices for the general population has found relatively consistent results between countries (11).

In conclusion, we found that there is limited availability of devices validated for use during pregnancy and these devices are more expensive than devices validated for the general population. To promote use of home BP devices specifically validated for use in pregnancy, it may be necessary to provide clear labelling at point-of-sale. Additionally, education about the need for specific validation of BP devices used during pregnancy may be beneficial for the patients, carers and healthcare providers.

## Sources of funding

NC is supported by a National Heart Foundation Australia Fellowship.

DSP is supported by a National Medical Research Council of Australia Fellowship (GNT2018077) and is an Honorary Future Leader Fellow of the Heart Foundation of Australia. The work was supported by New South Wales Cardiovascular Elite Postdoctoral Researcher Grant (H23/37663).

## Competing interests

No relevant disclosures.

## Data availability

The data that support the findings of this study are available from the corresponding author upon reasonable request.

## Author contributions

Conceptualisation, KS, NC, and DP.; methodology, KS, NC, and DP.; formal analysis, KS and HS.; writing—original draft preparation, KS.; writing—review and editing, KS, HS, NC and DP.; supervision, NC and DP.; project administration, KS. All authors have read and agreed to the final version of the manuscript.

---

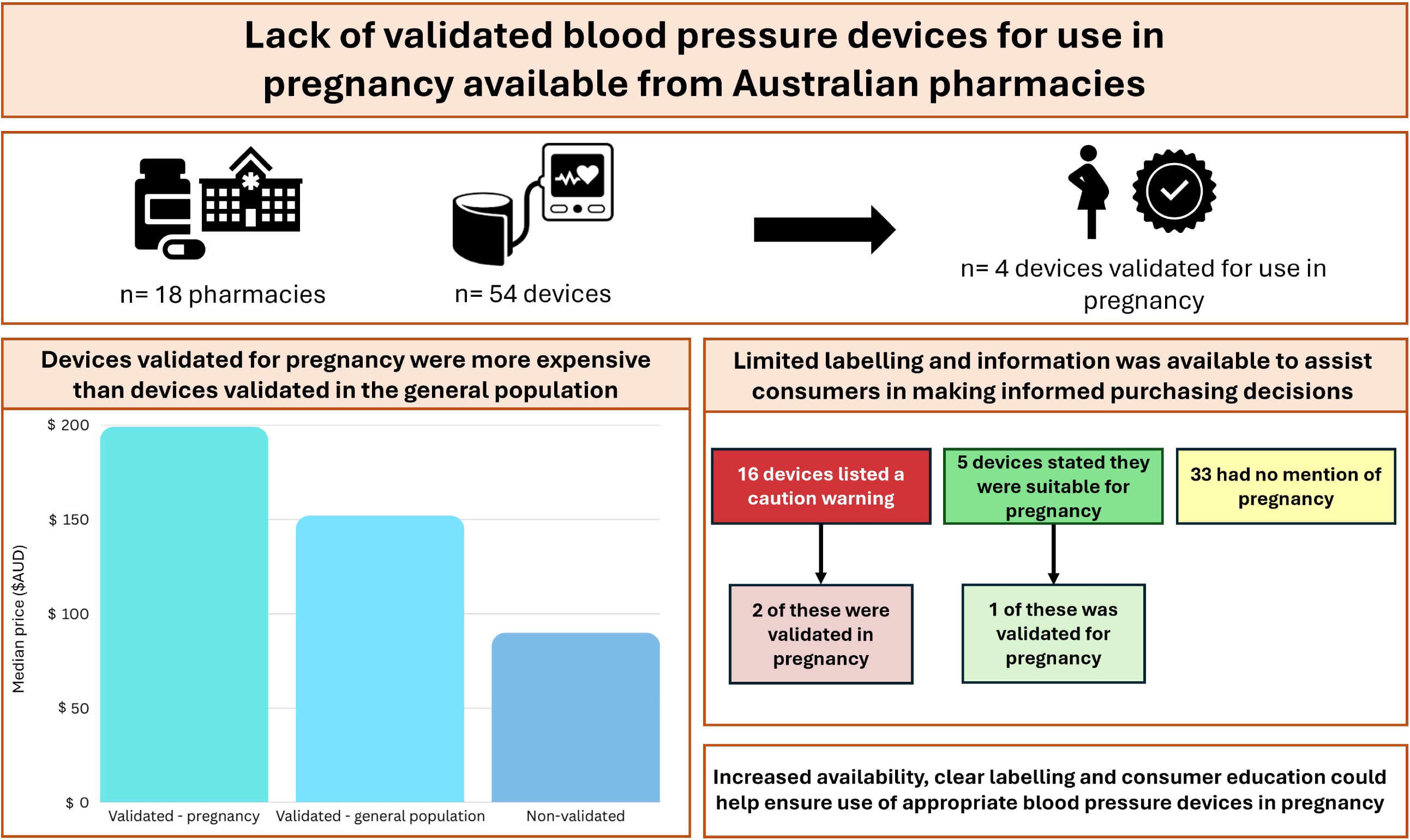

## Notes

### Competing Interest Statement

The authors have declared no competing interest.

## References

1. Society of Obstetric Medicine Australia and New Zealand. Hypertension in Pregnancy Guideline. Sydney; 2023.

2. Tucker KL, Sheppard JP, Stevens R, Bosworth HB, Bove A, Bray EP, et al. Self-monitoring of blood pressure in hypertension: A systematic review and individual patient data meta-analysis. PLoS Med. 2017;14(9):e1002389.

3. Imai Y, Obara T, Asamaya K, Ohkubo T. The reason why home blood pressure measurements are preferred over clinic or ambulatory blood pressure in Japan. Hypertens Res. 2013;36(8):661–72.

4. Picone DS, Deshpande RA, Schultz MG, Fonseca R, Campbell NRC, Delles C, et al. Nonvalidated Home Blood Pressure Devices Dominate the Online Marketplace in Australia. Hypertension. 2020;75(6):1593–9.

5. Hurrell A, Webster L, Chappell LC, Shennan AH. The assessment of blood pressure in pregnant women: pitfalls and novel approaches. Am J Obstet Gynecol. 2022;226(2s):S804-s18.

6. Bello NA, Woolley JJ, Cleary KL, Falzon L, Alpert BS, Oparil S, et al. Accuracy of Blood Pressure Measurement Devices in Pregnancy: A Systematic Review of Validation Studies. Hypertension. 2018;71(2):326–35.

7. Clapham E, Carmichael S, Picone DS, Schutte AE, Slater K, Stevens J, et al. How and why do Australians obtain blood pressure devices for use at home? A mixed-methods study. Hypertension Research [under review]. 2025:2025.02.27.24318446.

8. Stride BP. “BP Monitors” [Available from: https://www.stridebp.org/bp-monitors/.

9. Kalafat E, Benlioglu C, Thilaganathan B, Khalil A. Home blood pressure monitoring in the antenatal and postpartum period: A systematic review meta-analysis. Pregnancy Hypertension. 2020;19:44–51.

10. Tremonti C, Beddoe J, Brown MA. Reliability of home blood pressure monitoring devices in pregnancy. Pregnancy Hypertension: An International Journal of Women’s Cardiovascular Health. 2017;8:9–14.

11. Picone DS, Chapman N, Schultz MG, Schutte AE, Stergiou GS, Whelton PK, et al. Availability, Cost, and Consumer Ratings of Popular Nonvalidated vs Validated Blood Pressure-Measuring Devices Sold Online in 10 Countries. Jama. 2023;329(17):1514–6.

12. Schutte AE, Bennett B, Chow CK, Cloud GC, Doyle K, Girdis Z, et al. National Hypertension Taskforce of Australia: a roadmap to achieve 70% blood pressure control in Australia by 2030. Medical Journal of Australia. 2024;221(3):126–34.

